# Sustained Specific EBOV GP Immunogenicity Five-Years Post-Vaccination: Longitudinal Results from North Kivu and Equateur, Democratic Republic of the Congo

**DOI:** 10.64898/2026.05.19.26353493

**Authors:** Sydney Merritt, Nicole A. Hoff, Patrick K. Mukadi, Jean Paul Kompany, Megan Halbrook, Merly Tambu, Michael Beya, Handdy Kalengi, Victor Etuk, Teri Ann Wong, Jean Jacques Muyembe-Tamfum, J. Daniel Kelly, Didine K. Kaba, Lisa E. Hensley, Axel T. Lehrer, Jason Kindrachuk, Placide Mbala-Kingebeni, Anne W. Rimoin

## Abstract

Ebola virus disease (EVD), caused by the Ebola virus (EBOV), is characterized by high morbidity and mortality, with 16 distinct EVD outbreaks reported in the Democratic Republic of the Congo (DRC), alone. As part of the formal response to the 2018 outbreaks in Equateur and North Kivu provinces, a recombinant vesicular stomatitis virus-Zaire Ebolavirus envelope glycoprotein vaccine (rVSV-ZEBOV-GP) vaccine was deployed under emergency use. While clinical trials have evaluated vaccine safety and efficacy, there is a paucity of real-world data documenting antibody durability for longer periods post-vaccination. Here, we present serologic data from 1081 individuals in Beni, North Kivu (n = 599) and Mbandaka, Equateur (n = 482) who were vaccinated during the outbreaks–with samples from baseline up to five-years following vaccination. Using a multiplexed immunoassay, we show sustained anti-EBOV GP reactivity: at year-5 collection, 72% of individuals naïve at time of vaccination remained seroreactive to EBOV GP. Stratifying by site, antibody titers remained significantly elevated after baseline across all post-vaccination timepoints in both linear and logistic mixed-effects models. Pre-existing EBOV GP reactivity at baseline was the strongest independent predictor of antibody response in Mbandaka, associated with higher titers and greater odds of seropositivity (OR = 3.87, 95% CI: 2.50-6.01, p-value < 0.001), consistent with a boosting effect among previously exposed individuals. However, this was not replicated in Beni (OR: 0.66, 95% CI: 0.27-1.58, p-value = 0.348). In Mbandaka, among those recipients who reported receiving a booster dose, the odds of seroreactivity were 12.75-fold (p-value < 0.001) and 3.68-fold higher (p-value =0.04) at 4.2 and 5-years post-vaccination, respectively, in comparison to odds of reactivity at three weeks following administration of the initial dose. Occupational groups with zoonotic or community-level exposure had trending lower odds of seroreactivity relative to healthcare workers, most consistently in Beni. Ultimately, these data indicate that five years following administration of the rVSV-ZEBOV-GP vaccine, most vaccinated individuals retain detectable anti-EBOV GP antibodies. While no correlate of protection for EVD is established at this time, sustained IgG seroreactivity to EBOV GP may serve as a marker for future understandings of the durability of and variation in immune responses to this high-consequence pathogen.

## INTRODUCTION

*Orthoebolavirus zairense* (EBOV), the causative agent of Ebola virus disease (EVD), has caused infrequent outbreaks of human disease in Central and West Africa following the first identification of disease in humans in the Democratic Republic of the Congo (DRC). While outbreaks have primarily occurred in rural, forested regions, geographic expansion of EBOV into densely populated urban regions has resulted in devastating public health and economic consequences. EVD is a severe and often fatal disease characterized by multi-organ failure with potential hemorrhagic manifestations and an overall case fatality rate (CFR) of ∼50% (1, 2).

Since the first recorded EVD outbreak in 1976 near Yambuku village in Mongala province, the DRC has recorded 16 EVD outbreaks. This includes the second largest EVD outbreak ever recorded with 3,470 confirmed cases in North Kivu province from 2018 to 2020 (CFR 66%), and five subsequent outbreaks through 2022, with three more in North Kivu and two in Equateur province. Most recently, the 16th official EVD outbreak was declared in Bulape, Kasai Province in September 2025 (3). The increasing frequency and unpredictable nature of these outbreaks have resulted in expansive development of mitigation activities in the DRC which includes rapid diagnostic testing, reporting, and deployment of outbreak response support.

Beyond diagnostic deployment, the DRC Ministry of Health approved a compassionate use protocol for the use of rVSV-ZEBOV-GP (ERVEBO), a recombinant, replication competent vesicular stomatitis virus-based vaccine that expresses the glycoprotein (GP) of EBOV for deployment in 2017 (4). Preclinical data has demonstrated complete protection in EBOV-infected nonhuman primates when vaccinated 7-31 days prior to challenge as well as partial protection among animals vaccinated 3 days pre- to 24 hours-post exposure (5–7). Final results from *Ebola ça Suffit!*, a Phase III rVSV-ZEBOV-GP ring vaccination trial, indicated that no cases of EVD occurred 10 days or more after randomization among all immediately vaccinated contacts and contacts of contacts (8). Additionally, in a Phase II trial targeting safety and immunogenicity of a delayed booster dose of the vaccine (18 months post-primary immunization (9)) results indicated a several fold increase in the geometric mean ratio of antibody titers (GMT) among those who received a delayed booster. Specifically, the GMT of boosted individuals were 21-fold higher and 7-fold higher than those who received an additional non-booster dose at 19 months (1-month post-booster) and 36 months, respectively. Furthermore, monitoring of over 100,000 rVSV-ZEBOV-GP recipients between 2018 and 2020 has provided further evidence of high vaccine efficacy against EVD onset ten or more days post-vaccination (10). While these data have provided critical insights into the protective efficacy of rVSV-ZEBOV-GP, there is a paucity of data regarding longer term immunogenicity among vaccine recipients.

More recently, persistence and avidity of anti-EBOV GP antibodies has been reported at five years among a cohort of 168 vaccinees enrolled in Geneva, Switzerland and Lambaréné, Gabon (11). However, while results indicate an increase and plateau of elevated ZEBOV-GP immunoglobulin G (IgG) titers up to five years after vaccination, only participants (n = 84) from Switzerland were included in evaluations of antibody persistence. Additionally, recent cross-sectional studies among health care workers (HCW) from Kinshasa and Goma (n = 133) have demonstrated up to four-year immunogenicity of rVSV-ZEBOV-GP with strain-specific responses (12).

Given the sparsity of real-world data assessing post-rVSV-ZEBOV-GP immunogenicity up to five years after vaccination, we sought to evaluate longitudinal seroreactivity of two cohorts from regions of the DRC with recurrent EBOV outbreaks: Beni, North Kivu province, and Mbandaka, Equateur province. Early immunological data from Beni up to 6 months post-vaccination have indicated sustained anti-EBOV GP responses; initial analysis demonstrated that 87.2% of vaccinees had anti-GP IgG antibody titers 21 days post-vaccination, with antibody persistence out to 6 months post-vaccination among 95.6% of participants (13). Further, sex and age were found to be key predictors of seroreactivity with females and younger age being associated with higher antibody titers. In this study update, we sought to evaluate rVSV-EBOV immune durability up to 5 years post-vaccination among a complete study cohort at two geographically distinct sites (n = 1081 at baseline). Additionally, through this analysis, we endeavored to better understand driving factors associated with sustained immune responses and correlates of long-term protection in a setting with recurrent outbreak potential.

## METHODS

### Study design, setting and participants

We conducted a longitudinal cohort study of participants who had received the rVSV-ZEBOV-GP vaccine during the Equateur province and North Kivu province EVD outbreaks starting in 2018. Study inclusion for participants in the Beni, North Kivu cohort have been described previously (13) and these criteria for participants in Mbandaka, Equateur, were identical. Briefly, participants were enrolled at either site if they had received the rVSV-ZEBOV-GP vaccine under “compassionate use/expanded access” protocol using ring vaccination. In order to limit disruptions to the vaccination deployment led by the Expanded Program for Immunization (EPI) of the Ministry of Health and the World Health Organization (WHO), participants were not recruited for enrollment until after the 30-minute post-vaccine follow up period, nor were they offered compensation at the time of enrollment. Thus, those specific populations eligible for ring vaccination included: contacts, or contacts of contacts, of confirmed cases, healthcare workers and frontline workers in EVD affected areas. As children 1 year and above were eligible for vaccination, our study included those participants receiving the rVSV-ZEBOV-GP vaccine who were 1 year or older (with those under 18 assenting as appropriate). Pregnant women were included in the ring vaccination strategy, but were excluded from participation in this study, if pregnant at the time of enrollment.

In Equateur, enrollment began on June 7, 2018 through July 3, 2018 and in North Kivu, enrollment began on August 15, 2018 through August 29, 2018. Both cohorts were followed over the course of five years, with six collection periods in each province. Broadly, study visits included a baseline collection, and the six follow up visits from baseline vaccination: 21-28 days (V1), 6-7 months (V2), 1.5 years (V3, Beni only), 2.5 years (V4), 3.5 years (V5), 4.5 years and booster deployment (V6, Mbandaka only) and 5.2 years (V7) **(Figure 1)**.

**Figure 1.**
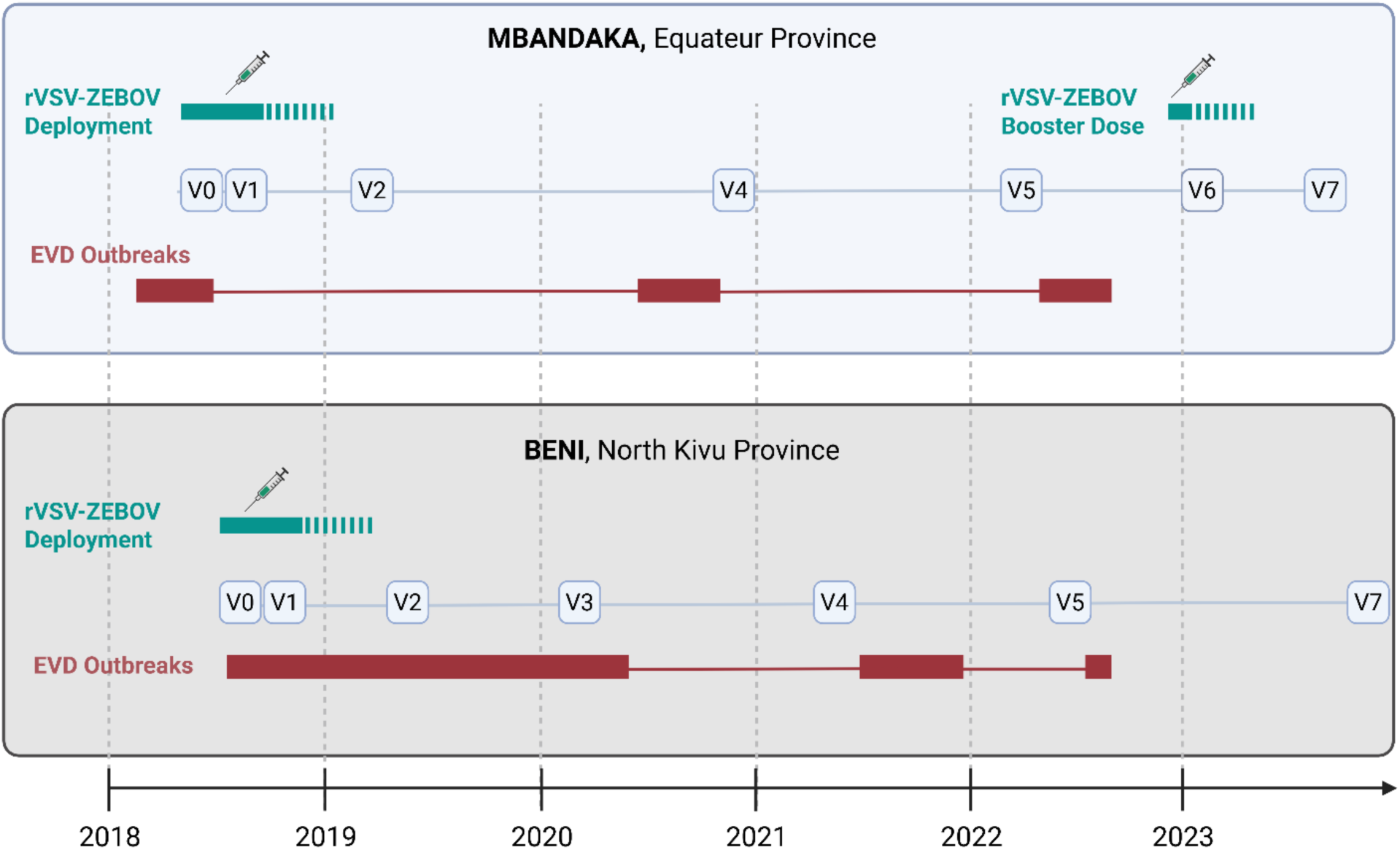
Timeline of Serum Sample Collection in Mbandaka and Beni, Democratic Republic of the Congo. Corresponding reported Ebola virus disease (EVD) outbreaks in both respective sites are presented in red. Vaccination deployment was exclusively directed under a compassionate use protocol for use during the 2018 EVD outbreaks of Mbandaka and Beni.

At each study visit, participants completed an interviewer-led questionnaire capturing sociodemographic data, zoonotic exposure data, vaccination and booster dose data, and vaccine knowledge, attitudes and behaviors. Participants also completed a short physical assessment which included height, weight, blood pressure and heart rate, and provided a blood specimen. Procedures following blood collection have been described previously (13). In short, after collection, all blood samples were centrifuged, and serum samples heat inactivated for transport and storage; 1mL serum aliquots were stored at -20C in field laboratories and transferred to -80C storage at the National Institute for Biomedical Research (INRB) in Kinshasa.

### Serologic Assessments

Serologic testing was completed using a pan-filovirus multiplex immunoassay (MIA) which included purified recombinant EBOV GP, EBOV nucleoprotein (NP), and EBOV viral protein 40 (VP40) targets(14, 15). Additionally, this MIA included bovine serum albumin (BSA)-coupled beads as an indicator of background reactivity. The EBOV GP reactivity component of this has been previously compared to the gold-standard ELISA, Filovirus Animal Non-clinical Group, or FANG assay, with strong correlation between detected antibody reactivity (16). MIA methods have been previously described (17); in brief, samples were diluted to 1:100 using 2 µL of serum and run in duplicate with an output of median fluorescent intensity (MFI) for each antigen. Any technical duplicates with a difference in MFI outputs > 20% for any antigen target were flagged for retesting. Before analysis, detected anti-BSA background MFI was subtracted from the mean MFI for each filoviral antigen for each individual. Seroreactivity thresholds have been previously determined as the mean MFI plus three standard deviations of a filovirus-naive Kinshasa cohort after BSA-adjustment (n = 379, (14)).

### Statistical Analyses

Here, we report the log-transformed mean BSA-adjusted MFI values for each antigen. As the multiplexed serologic assay at a single dilution and without a standard curve is not capable of quantifying antibody concentrations, relative changes in antibody reactivity since baseline are reported, allowing determination of a binary seroreactivity status for each subject at each time point. Baseline reactivity was established using only the antigen-specific cut-off of 2589 MFI for EBOV GP. Individuals were considered to have seroconverted (“seroreactive”) at follow-up visits if detected MFI values were at least four-fold greater than the corresponding baseline visit MFI (18), and above the pre-established threshold for EBOV GP reactivity. These are conservative criteria for seroreactivity, as those moderately reactive (but below the MFI) cut off at baseline may not mount a full four-fold increase post-vaccination. For each visit, follow-up rates were calculated as percentages of the total participants enrolled at baseline. Univariate distributions of key demographic characteristics were presented for both cohorts at baseline. Wald 95% confidence interval (95% CI) estimates are reported as applicable.

### Model Fitting

To assess the univariate predictors of anti-EBOV GP seroreactivity, participant age, sex, education, occupation (grouped by possible exposure route), site, and baseline reactivity were assessed at each follow up time-point using linear mixed effect models. Site was not included as a covariate in any multivariate models due to marked differences in antibody reactivity patterns over the five-year follow-up **(Figure 2).** Given these site-based differences in baseline reactivity, booster dose administration, follow-up visit timing, and MFI trajectories over time, all analyses were stratified by study site (Beni or Mbandaka).

**Figure 2.**
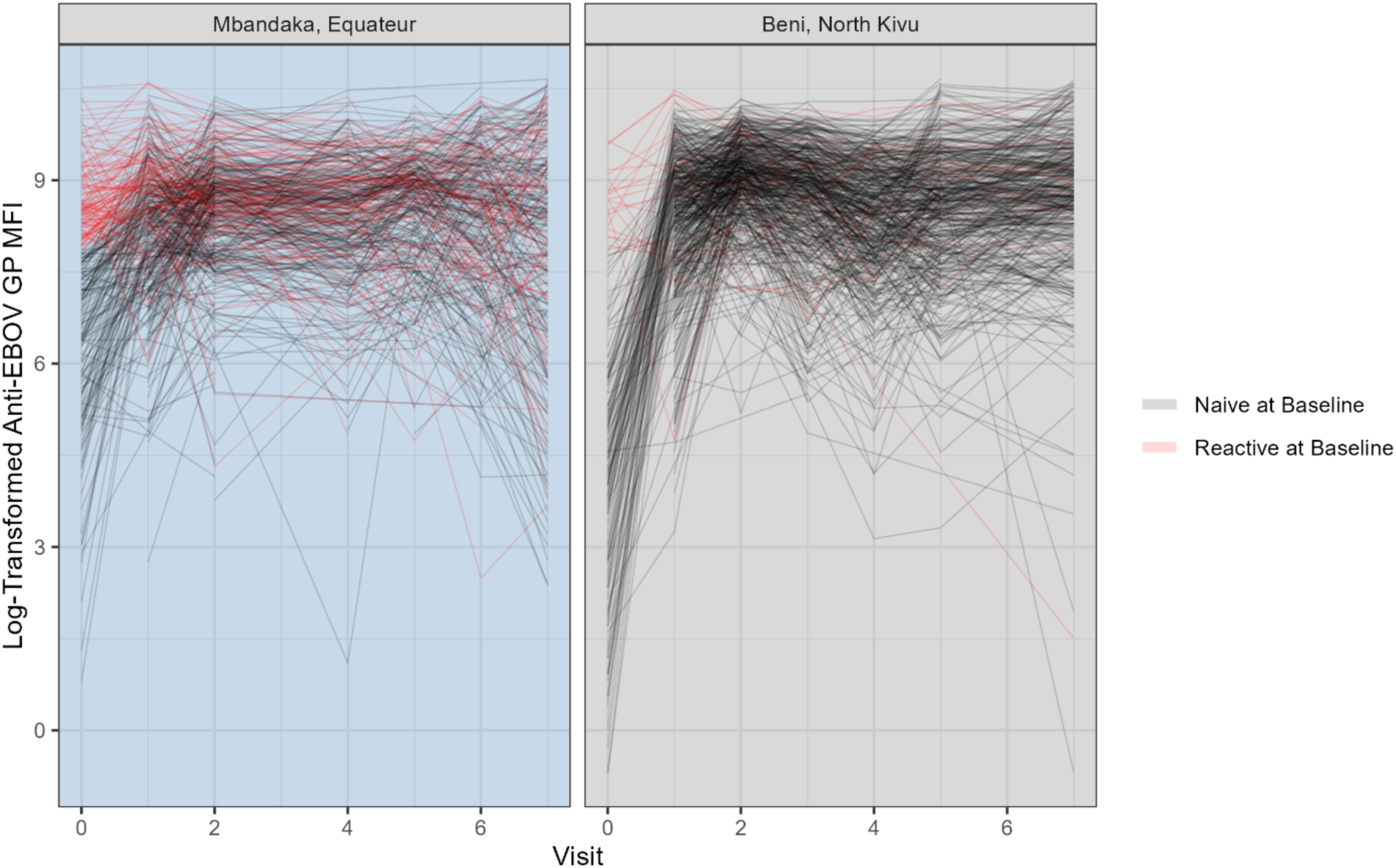
Individual Trajectories of log-transformed anti-EBOV GP MFI by site over seven visits. Those respondents considered seroreactive to EBOV GP at baseline are denoted in red.

Mixed effects models were specifically selected for this analysis due to the repeated measures of individual vaccinees, and inherent correlation between serological measures of the same individuals. Selected predictors were then included in a multivariate linear mixed effects model: sex, age, occupation by potential exposure route, education level (as a proxy for socio-economic status), self-reported booster dose reception (at V6 in Mbandaka only) and reactivity at baseline. Model diagnostics indicated that linear model fit was improved with the inclusion of both visit and unique identifier as random effects, both random slope and intercept variables. Furthermore, high collinearity (r^2^ = 0.98) between time since baseline in days and visit number indicated interchangeable use of either variable as the time component; visit was selected as the time indicator for all modeling approaches. Only complete cases (individuals with all included covariates) were assessed in model fits to improve model stability, and an interaction term of visit and baseline reactivity was included to determine whether pre-existing reactivity to EBOV GP prior to vaccination influenced later vaccine-induced anti-EBOV GP reactivity and sustained antibody durability over the time of follow-up. Models were run for each site stratum were identical bar the inclusion of a booster dose-visit interaction term for Mbandaka. All models treated participants as unique subjects with additive random effects and compound symmetry correlation structures, and all predictors as fixed effects. Autoregressive correlation structures were evaluated and marginally improved information criterion outputs, yet due to the unequal spacing of visits and minimal improvement of model fit, no change to the compound symmetry structure was indicated.

Given the binary seroreactivity output of the MIA as established by the MFI cut off and four-fold change approach, we next repeated these assessments of antibody durability predictors using a multivariate logistic mixed effects model. No visit and baseline reactivity interaction term was included for this logistic mixed effect model approach as baseline reactivity was determined with the same cut-point criterion as the binary outcome of seroreactivity.

Analyses were conducted in SAS (version 9.4, Cary, NC) and R (version 4.5.2, R Core Team, 2026).

### Ethical Considerations

All study activities were conducted under approval from appropriate institutional review boards: University of California, Los Angeles IRB No. IRB-16-001346 and IRB-20-001091, and Kinshasa School of Public Health IRB No. ESPCE0222017. Secondary analysis was conducted using a de-identified dataset with serologic results from all consenting individuals. Additionally, the study was approved by the Scientific Committee for Ebola Research during an outbreak at the INRB under the Ministry of Health. Prior to data or serum collection, participants signed or marked the approved informed consent form, and parents or guardians provided this consent on behalf of all child participants, while adolescents aged 7 to 17 provided assent as appropriate.

## RESULTS

Over the course of the five-year follow-up in Beni and Mbandaka, 1081 unique individuals who had received the rVSV-ZEBOV-GP vaccine were recruited for participation **(Table 1)**. Notably, follow-up rates at both sites were greater than 60% for all visits except V6, Mbandaka, due to specific recruitment and follow up of those respondents who had likely received a booster dose.

**Table 1.** Study Population Distribution by Site and Months Since Vaccination.

| Visit No. | Time Since Vaccination | Mbandaka, EQT |  |  | Beni, NKI |  |  | Total N |
| --- | --- | --- | --- | --- | --- | --- | --- | --- |
|  |  | N | % Follow Up | Collection Date | N | % Follow Up | Collection Date |  |
| V0 | Baseline | 482 | -- | Jun 2018 | 599 | -- | Aug 2018 | 1081 |
| V1 | 21 Days | 467 | 96.9 | Jul 2018 | 541 | 90.3 | Sep 2018 | 1008 |
| V2 | 6 Months | 471 | 97.7 | Feb 2019 | 501 | 83.6 | Apr 2019 | 918 |
| V3 | 1.5 Years |  |  |  | 467 | 78.0 | Feb 2020 | 467 |
| V4 | 2.5 Years | 355 | 73.7 | Dec 2020 | 373 | 62.3 | Apr 2021 | 728 |
| V5 | 3.5 Years | 329 | 68.3 | Mar 2022 | 391 | 65.3 | May 2022 | 720 |
| V6 | 4.2 Years + Booster | 198 | 41.1 | Feb 2023 |  |  |  | 198 |
| V7 | 5+ Years | 304 | 63.1 | Aug 2023 | 425 | 71.0 | Dec 2023 | 729 |

At baseline, most participants were male (66.1%) and thirty years old or older **(Table 2)**. Approximately one-third of the vaccinated participants listed health care professions as their primary occupation in both Beni (34.9%) and Mbandaka (33.2%). Beyond occupational status, 15.0% of the overall population sample were reactive to EBOV GP at Visit 0, prior to receiving the vaccine; notably, this differed significantly by site, with 28.8% of participants in Mbandaka reactive at baseline, compared to 3.8% of participants in Beni. In Mbandaka alone, 52 participants at baseline (10.8%) later self-reported reception of a booster dose of r-VSV-ZEBOV-GP four years and two months following primary vaccination.

**Table 2.** Population Demographic Characteristics by Site at Baseline, Visit 0. All participants were enrolled if they had received the rVSV-ZEBOV vaccine as part of the 2018 response to the two separate Ebolavirus outbreaks in Beni, North Kivu Province, and Mbandaka, Equateur Province, in 2018. Study population has been stratified by site to reflect key differences in outbreak settings, booster dose administration, and baseline seroreactivity detected in the two selected provinces.

| Sample Characteristic | All<br>n = 1081 |  | Mbandaka, EQT<br>n = 482 |  | Beni, NKI<br>n = 599 |  |
| --- | --- | --- | --- | --- | --- | --- |
|  | N | % | N | % | N | % |
| **EBOV GP Seroreactive at Baseline** |  |  |  |  |  |  |
| No | 919 | 85.0 | 343 | 71.2 | 576 | 96.2 |
| Yes | 162 | 15.0 | 139 | 28.8 | 23 | 3.8 |
| **Self-Reported Booster Dose at V6** |  |  |  |  |  |  |
| No |  |  | 430 | 89.2 |  |  |
| Yes (Booster = 1) |  |  | 52 | 10.8 |  |  |
| **Sex** |  |  |  |  |  |  |
| Male | 715 | 66.1 | 332 | 68.9 | 383 | 63.9 |
| Female | 367 | 34.0 | 150 | 31.1 | 217 | 36.2 |
| **Age Category** |  |  |  |  |  |  |
| 12-19 | 97 | 9.0 | 40 | 8.3 | 57 | 9.5 |
| 20-29 | 229 | 21.2 | 69 | 14.3 | 160 | 26.7 |
| 30-39 | 334 | 30.9 | 138 | 28.6 | 196 | 32.7 |
| 40-49 | 239 | 22.1 | 130 | 27.0 | 109 | 18.2 |
| 50+ | 182 | 16.8 | 105 | 21.8 | 77 | 12.9 |
| **Civil Status** |  |  |  |  |  |  |
| Single | 370 | 34.2 | 130 | 27.0 | 240 | 40.1 |
| Married or Living Together as Married | 681 | 63.0 | 339 | 70.3 | 342 | 57.1 |
| Divorced, Separated or Widowed | 27 | 2.5 | 13 | 2.7 | 14 | 2.3 |
| Unknown | 0 | 0.0 | 3 | 0.6 | 3 | 0.5 |
| **Educational Level** |  |  |  |  |  |  |
| No Formal Education | 28 | 2.6 | 7 | 1.5 | 21 | 3.5 |
| Any Primary School or Apprenticeship | 319 | 29.5 | 74 | 15.4 | 245 | 40.9 |
| Completed Secondary School | 291 | 26.9 | 136 | 28.2 | 155 | 25.9 |
| Post-Secondary Education | 442 | 40.9 | 264 | 54.8 | 178 | 29.7 |
| Refused | 1 | 0.1 | 1 | 0.2 | 0 | 0.0 |
| **Occupation (by Exposure Route)** |  |  |  |  |  |  |
| Health Care Worker | 369 | 34.1 | 160 | 33.2 | 209 | 34.9 |
| Zoonotic Exposure (Farmers, Hunters) | 209 | 19.3 | 81 | 16.8 | 128 | 21.4 |
| Human to Human Exposure (Merchants, Teachers, Politicians) | 154 | 14.2 | 75 | 15.6 | 79 | 13.2 |
| Other | 337 | 31.2 | 160 | 33.2 | 177 | 29.5 |
| Refused | 12 | 1.1 | 6 | 1.2 | 6 | 1.0 |

Over the course of follow up, the majority of all participants remained seroreactive to EBOV GP following vaccination. On the relative scale of log-MFI, only the baseline visit median MFI output was below the set cut off **(Figure 3)** with highest anti-EBOV GP reactivity detected at Visit 2, or six-months post-vaccination.

**Figure 3.**
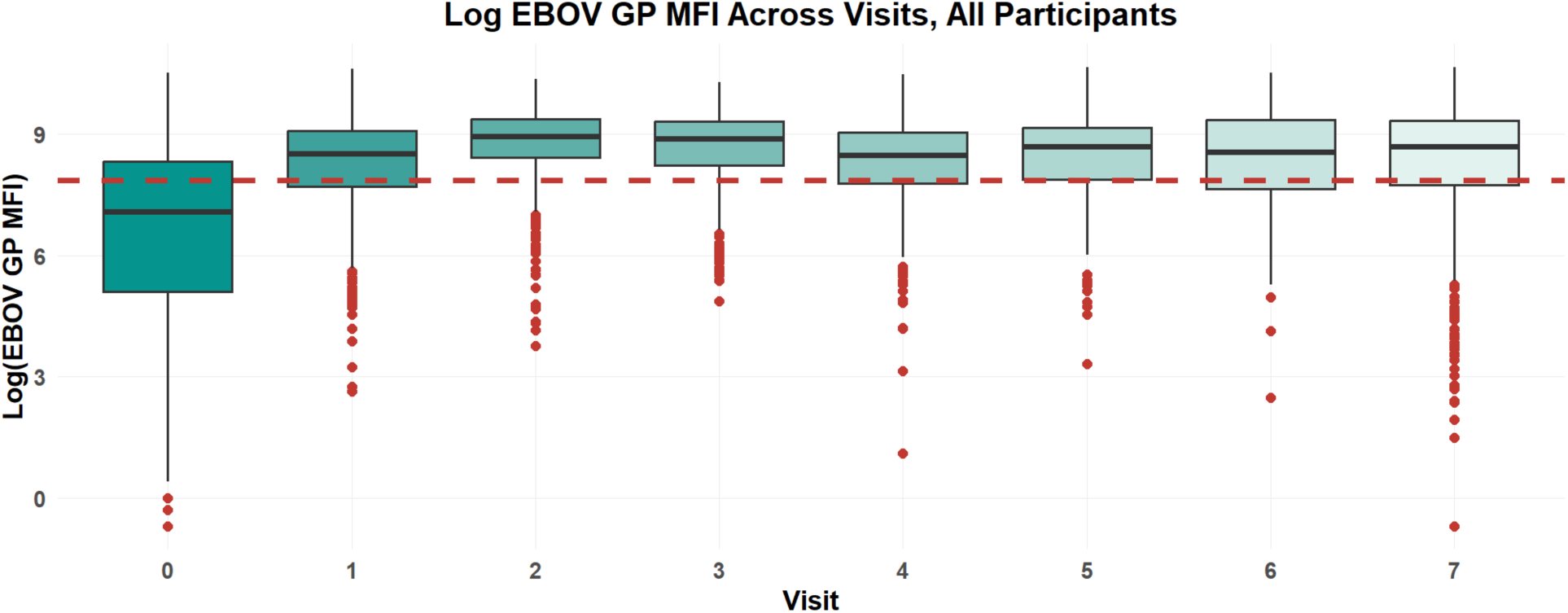
Log-Anti-EBOV GP MFI across all visits. Cut off as indicated by Kinshasa Naïve Population Mean MFI + 3 standard deviations.

When binarized, at Visit 7, or five-years post-vaccination, 71% of all participants were reactive to EBOV GP **(Figure 4)**. By site, 81% and 57% of Beni and Mbandaka participants, respectively were seroreactive at Visit 7. Stratified by sex, 75% and 68% of female and male participants remained seroreactive at five-years. Notably, among those seroreactive at baseline (“No”, naïve at baseline), 65% were seroreactive at last collection, as opposed to those EBOV GP-naïve at baseline, where 72% remained seroreactive. At six-months post-vaccination, we observed no difference in the proportion of seroreactivity to EBOV GP by baseline reactivity status – both groups were 84% seroreactive at Visit 2.

**Figure 4.**
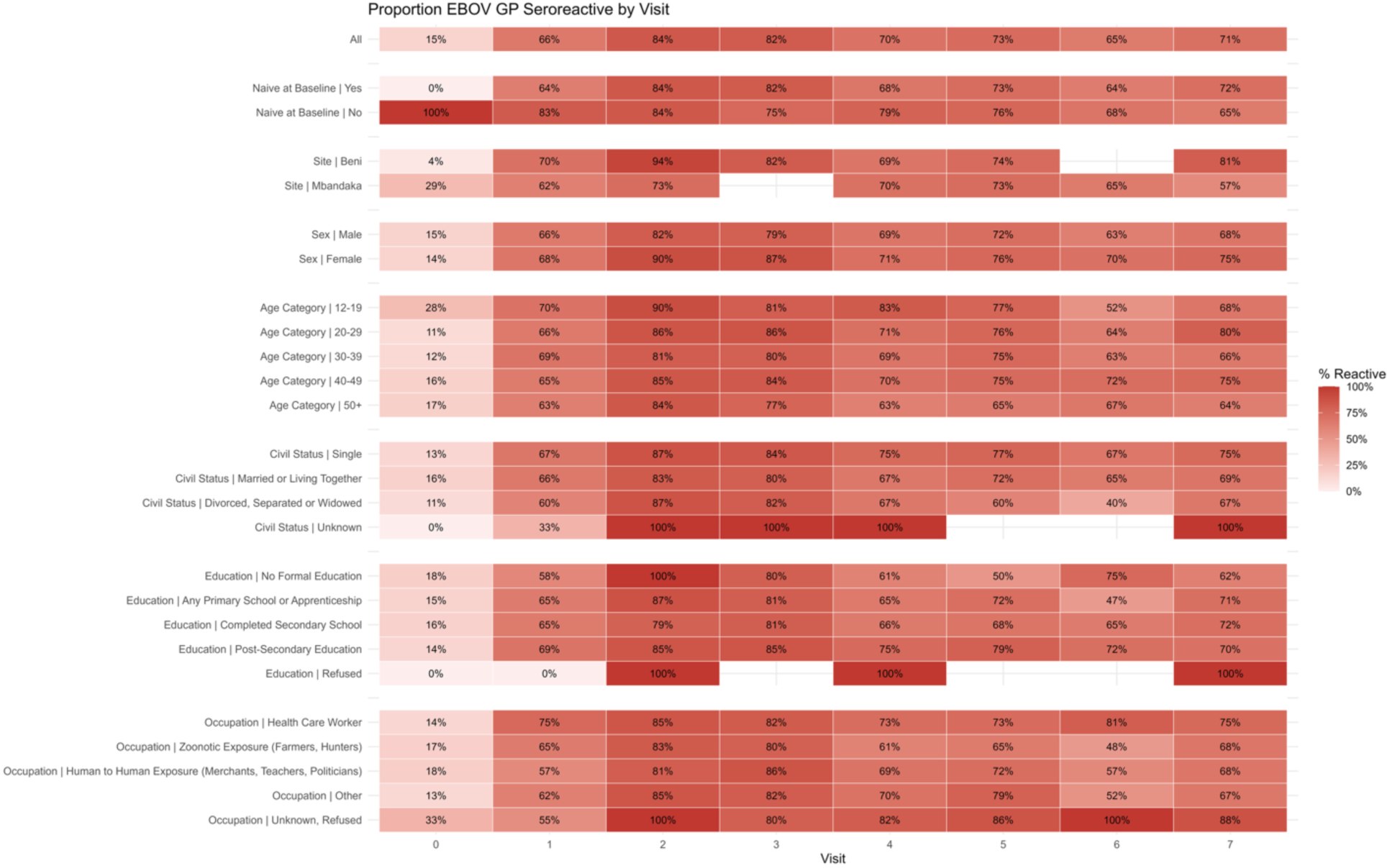
Proportion of EBOV GP Seroreactivity individuals by Visit. Seroreactivity was determined as those with log-MFI values greater than the EBOV GP-specific cut off of log (2589 MFI) at baseline Visit 0, and for all subsequent values, if detected MFI values were both greater than the established cut off and greater than four-fold the baseline MFI of each participant.

Excluding those considered seroreactive at baseline, the greatest mean peak log-MFI values were detected at Visit 2, with 8.8 (95% CI: 8.7-8.9) overall, and 8.3 (95% CI: 8.2-8.5) and 9.0 (95% CI: 9.0-9.1) in Mbandaka and Beni, respectively **(Supplementary Table 1-2)**. When dichotomized, these trends in seroreactivity hold among an entirely naïve-at-baseline population. At six-months, 61.5% and 93.6% of participants in Beni and Mbandaka, respectively, were considered reactive to EBOV GP. By Visit 7, these proportions of seroreactive were 51.5% and 79.9% at the two sites, indicating some decline, yet a sustained antibody response to EBOV GP following vaccination with rVSV-ZEBOV-GP.

Visit 6 collection in Mbandaka was specifically planned to coincide with administration of a booster dose of the rVSV-ZEBOV-GP vaccine. Of those 198 individuals enrolled in February 2023, 46 (23.2%) reported having received the booster dose. At Visit 6 collection, of these 46 individuals, 39/46 were seroreactive to EBOV GP (84.7%), and at Visit 7, of those 44 booster dose recipients recruited, 29/44 maintained seroreactivity to EBOV GP (65.9%).

### Linear Mixed Effect Model

Stratifying by site (Mbandaka, n = 462, Beni, n = 573), linear mixed effects models were fitted to identify predictors of log-transformed anti-EBOV GP antibody titer change over time, with random intercepts and random slopes to account for individual-level variability in baseline reactivity and immunodynamics **(Table 3)**. Visit 1, or 21-days post-vaccination, was set as the reference group for detected antibody change over time, with the assumption that all vaccinated individuals at Visit 0 would have a detected anti-EBOV GP response three weeks following vaccination.

**Table 3.**
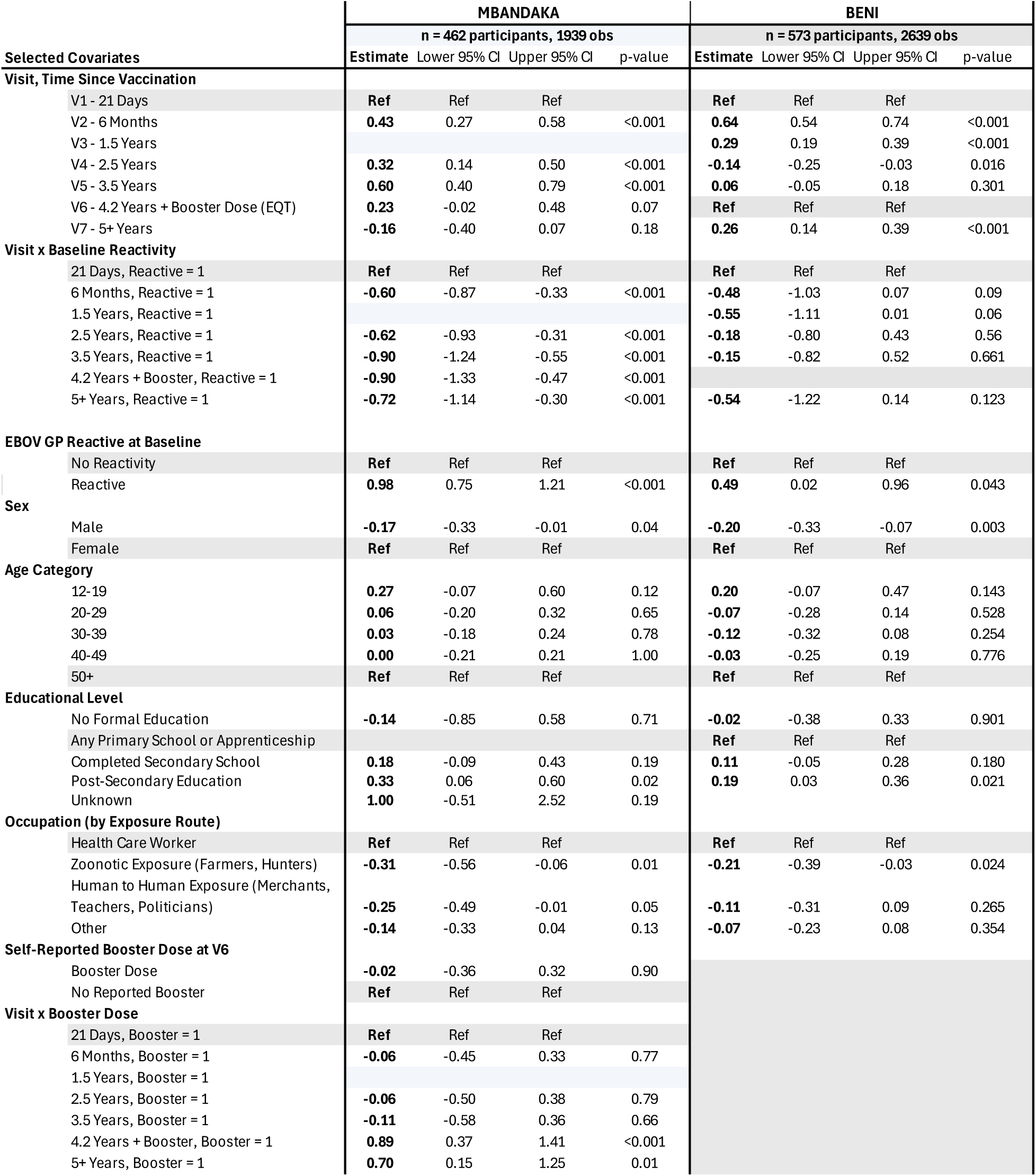
Linear mixed-effects model estimates of predictors of EBOV GP antibody titers over time, Mbandaka (n = 462) and Beni (n = 573) cohorts.

At both sites, adjusting for age, sex, education level, occupation, and the potential interaction of visit time and baseline reactivity, antibody titers were stable post-day 21 following vaccination with titers peaking in Mbandaka at Visit 5, 3.5 years post-vaccination (β = 0.60, 95% CI: 0.40–0.79, p-value < 0.001), and at six-months post-vaccination in Beni (β = 0.64, 95% CI: 0.54-0.73, p-value < 0.001). In Mbandaka, we observe a modest decline in anti-EBOV GP antibody titers at five years from Day-21 reactivity, yet still well above the baseline titers (β = -0.16, 95% CI: - 0.40-0.07, p-value = 0.18). Interestingly, at five-years post-vaccination in Beni, detected antibody titers were significantly elevated beyond those detected at day-21 (β = 0.26, 95% CI: 0.14 -0.39, p-value <0.001), which may be indicative of additional exposure beyond vaccination.

In Mbandaka, we observe a significant positive interaction effect of the booster dose and time since vaccination, with estimated antibody titers diverging at Visit 6 and Visit 7 (Visit 6: β = 0.89, p-value <0.001; Visit 7: β = 0.70, p-value = 0.01). These results indicate an additional effect of the booster dose on detected antibody titers following administration at Visit 6, after adjusting for age, sex, occupation, education and baseline reactivity of participants.

Age appeared to have a limited independent influence on antibody titers with only a slightly higher antibody titers in participants aged 12-19 at baseline in Mbandaka observed (β = 0.27, 95% CI: -0.07-0.60). Notably, male sex was associated with marginally lower antibody titers in both Mbandaka (β = -0.17, 95% CI: -0.33 to -0.01) and Beni (β = -0.20, 95% CI: -0.33 to -0.07, p-value = 0.003). A significant interaction between baseline reactivity and time since vaccination was present at both sites, with negative interaction term estimates across all visits at all sites relative to baseline reactivity. For example, at the six-month visit in Mbandaka and Beni, interaction term estimates were −0.60 (95% CI: -0.87 to -0.33) and -0.48 (95% CI: -1.03 to 0.07), suggesting that vaccine-induced fold-increase in antibody titers was substantially lower among those with pre-existing evidence of EBOV GP exposure as indicated by binary seroreactivity measures. These results were more pronounced among Mbandaka participants than among those from Beni. However, looking at baseline-reactivity alone, pre-existing reactivity to EBOV GP at the time of vaccination was positively associated with antibody titer increases at both sites (Mbandaka: β = 0.98, 95% CI: 0.75-1.21; Beni: β = 0.49, 95% CI: 0.02- 0.96).

### Logistic Mixed Effect Model

Mixed effects logistic regression models were fit separately for Beni (n = 593) and Mbandaka (n = 469) to assess predictors of EBOV GP seroreactivity following vaccination with rVSV-ZEBOV-GP. Identical random intercept and slope structures, identical reference group structures and the same covariates were included, as in the linear mixed effects model above. However, no baseline reactivity and time interaction term was included, as this baseline anti-EBOV GP seroreactivity was determined using the same cut off approach as the primary outcome in this model **(Table 4)**.

**Table 4.** Logistic mixed-effects model odds ratio estimates of predictors of EBOV GP seroreactivity over time, Mbandaka (n = 469) and Beni (n = 593) cohorts.

| Selected Covariates | MBANDAKA |  |  |  | BENI |  |  |  |
| --- | --- | --- | --- | --- | --- | --- | --- | --- |
|  | n = 469 participants, 2041 obs |  |  |  | n = 593 participants, 3264 obs |  |  |  |
|  | Odds Ratio | Lower 95% CI | Upper 95% CI | p-value | Odds Ratio | Lower 95% CI | Upper 95% CI | p-value |
| **Visit, Time Since Vaccination** |  |  |  |  |  |  |  |  |
| V1 - 21 Days | Ref | Ref | Ref |  | Ref | Ref | Ref |  |
| V2 - 6 Months | 1.98 | 1.39 | 2.82 | <0.001 | 9.57 | 6.11 | 14.97 | <0.001 |
| V3 - 1.5 Years |  |  |  |  | 3.33 | 2.27 | 4.88 | <0.001 |
| V4 - 2.5 Years | 1.92 | 1.27 | 2.89 | 0.00 | 1.32 | 0.88 | 1.98 | 0.185 |
| V5 - 3.5 Years | 2.87 | 1.77 | 4.65 | <0.001 | 2.66 | 1.62 | 4.37 | <0.001 |
| V6 - 4.2 Years + Booster | 1.09 | 0.61 | 1.95 | 0.76 |  |  |  |  |
| V7 - 5+ Years | 0.80 | 0.49 | 1.31 | 0.38 | 9.28 | 4.45 | 19.35 | <0.001 |
| **EBOV GP Reactive at Baseline** |  |  |  |  |  |  |  |  |
| No Reactivity | Ref | Ref | Ref |  | Ref | Ref | Ref |  |
| Reactive | 3.87 | 2.50 | 6.01 | <0.001 | 0.66 | 0.27 | 1.58 | 0.348 |
| **Sex** |  |  |  |  |  |  |  |  |
| Male | 0.73 | 0.49 | 1.09 | 0.12 | 0.60 | 0.42 | 0.86 | 0.006 |
| Female | Ref | Ref | Ref |  | Ref | Ref | Ref |  |
| **Age Category** |  |  |  |  |  |  |  |  |
| 12-19 | 1.39 | 0.62 | 3.12 | 0.43 | 1.79 | 0.85 | 3.79 | 0.126 |
| 20-29 | 0.96 | 0.52 | 1.79 | 0.90 | 1.04 | 0.59 | 1.82 | 0.903 |
| 30-39 | 1.02 | 0.61 | 1.69 | 0.95 | 0.93 | 0.55 | 1.58 | 0.793 |
| 40-49 | 0.95 | 0.57 | 1.58 | 0.85 | 1.05 | 0.58 | 1.89 | 0.875 |
| 50+ | Ref | Ref | Ref |  | Ref | Ref | Ref |  |
| **Educational Level** |  |  |  |  |  |  |  |  |
| No Formal Education | 0.68 | 0.11 | 4.26 | 0.68 | 0.91 | 0.37 | 2.27 | 0.847 |
| Any Primary School or Apprenticeship |  |  |  |  | Ref | Ref | Ref |  |
| Completed Secondary School | 1.14 | 0.62 | 2.12 | 0.68 | 1.17 | 0.75 | 1.82 | 0.481 |
| Post-Secondary Education | 2.39 | 1.26 | 4.53 | 0.01 | 1.67 | 1.07 | 2.63 | 0.026 |
| Unknown | 0.95 | 0.03 | 35.34 | 0.98 | -- | -- | -- | -- |
| **Occupation (by Exposure Route)** |  |  |  |  |  |  |  |  |
| Health Care Worker | Ref | Ref |  |  | Ref | Ref | Ref |  |
| Zoonotic Exposure (Farmers, Hunters) | 0.68 | 0.37 | 1.23 | 0.20 | 0.61 | 0.38 | 1.00 | 0.050 |
| Human to Human Exposure (Merchants, Teachers, Politicians) | 0.88 | 0.49 | 1.57 | 0.66 | 0.59 | 0.35 | 1.02 | 0.056 |
| Other | 0.79 | 0.50 | 1.24 | 0.30 | 0.62 | 0.40 | 0.95 | 0.028 |
| **Self-Reported Booster Dose at V6** |  |  |  |  |  |  |  |  |
| Booster Dose | 0.66 | 0.32 | 1.38 | 0.27 |  |  |  |  |
| No Reported Booster | Ref | Ref | Ref |  |  |  |  |  |
| **Visit x Booster Dose** |  |  |  |  |  |  |  |  |
| 21 Days, Booster = 1 | Ref | Ref | Ref |  |  |  |  |  |
| 6 Months, Booster = 1 | 0.96 | 0.35 | 2.60 | 0.93 |  |  |  |  |
| 1.5 Years, Booster = 1 |  |  |  |  |  |  |  |  |
| 2.5 Years, Booster = 1 | 1.27 | 0.42 | 3.86 | 0.68 |  |  |  |  |
| 3.5 Years, Booster = 1 | 0.81 | 0.25 | 2.57 | 0.72 |  |  |  |  |
| 4.2 Years + Booster, Booster = 1 | 12.75 | 3.01 | 54.04 | <0.001 |  |  |  |  |
| 5+ Years, Booster = 1 | 3.68 | 1.04 | 12.97 | 0.04 |  |  |  |  |

Adjusting for age, sex, education, baseline reactivity, booster dose (Mbandaka only) and occupation, the odds of seroreactivity increased markedly following vaccination in comparison to day-21 reactivity at both sites. In Mbandaka, six-months post-vaccination the odds of seroreactivity were 1.98 times that at three-weeks (95% CI: 1.39-2.82). Five years later, the odds of seroreactivity were not significantly reduced in relation to day-21 odds (OR = 0.80, 95% CI: 0.49-1.31, p-value = 0.38). Results were more pronounced in Beni, with the odds of seroreactivity at six-months post vaccination 9.57 times that of seroreactivity at day-21 (95% CI6.11-14.97, p-value < 0.001) and even more pronounced at the five-year post-vaccination time point (OR = 9.28, 95% CI 4.45-19.35). Increased odds of seroreactivity were maintained across sites and all time-points of follow-up relative to day-21 post-vaccination, except for Visit 7 Mbandaka.

However, interaction terms in the Mbandaka-only model suggest a significant divergence in the odds of seroreactivity following the administration of a booster dose at Visit 6. Aligned with results from the linear mixed effects model above, among those recipients who reported receiving a booster dose, the odds of seroreactivity were 12.75-fold (95% CI: 3.10-54.04, p-value < 0.001)and 3.68-fold higher (95% CI: 1.04-12.97, p-value =0.04) at 4.2 and 5-years post-vaccination, respectively, in comparison to odds of reactivity at three weeks following administration of the initial dose, after adjusting for age, sex, education, occupation and baseline reactivity.

Pre-existing seroreactivity to EBOV GP was significantly associated with an almost four-fold increase in the odds of seroreactivity in Mbandaka (OR = 3.87, 95% CI: 2.50-6.01, p-value < 0.001). However, this association was not observed in Beni, where the odds of seroreactivity among those reactive at baseline was not significantly associated with overall odds of seroreactivity post-vaccination (OR: 0.66, 95% CI: 0.27-1.58, p-value = 0.348). Male sex was associated with modestly lower odds of seroreactivity in both sites, with a more pronounced effect in Beni than Mbandaka. Similar to change in antibody titers, no statistically significant associations were observed between age category and seroreactivity at either site. Relative to healthcare workers, participants with occupations putting them at risk of zoonotic exposure had lower odds of seropositivity in Beni (OR = 0.61, 95% CI: 0.38-1.00) but not significantly so in Mbandaka (OR = 0.74, 95% CI: 0.41-1.33).

## DISCUSSION

Overall, we demonstrate sustained anti-EBOV GP seroreactivity up to five years post-rVSV-ZEBOV-GP vaccination among a cohort of 1081 individuals in two geographically distinct EBOV-endemic regions of the Democratic Republic of the Congo: Beni, North Kivu Province and Mbandaka, Equateur province. In this study, we describe population-level proportions of seroreactivity over five-years following emergency deployment of the rVSV-ZEBOV-GP vaccine, as well as individual-level predictors of EBOV GP antibody responses, both relative changes in antibody titers, as well as the odds of the binary outcome of seroreactivity to EBOV glycoprotein antigen. These findings expand upon prior clinical and observational findings by demonstrating durable antibody responses in high-risk populations, providing one of the largest and longest real-world evaluations of this vaccine to date (8, 19–21).

Across both mixed effect model approaches, vaccination was associated with sustained and durable antibody responses to EBOV GP. Pre-existing baseline reactivity, indicative of prior exposure to EBOV or a related pathogen and probable immune priming, was significantly associated with antibody response in Mbandaka, Equateur Province. This finding is aligned with work in other vaccine-elicited antibody fields where prior exposure to the pathogen, or pre-existing pathogen-specific titers have been shown to be the strongest predictors of post-vaccination antibody titers (22, 23). Yet, interaction terms indicate an attenuation of vaccine-induced fold-change of antibody responses; these data may suggest an immunological ceiling—those previously exposed could not be “boosted” at the same magnitude as those participants who were immuno-naïve to EBOV GP at the time of vaccination—yet additional data is needed to further support this claim. Furthermore, samples in this assay were run at a single dilution, where a saturating condition of the assay itself may create an artificial ceiling of detection. Future analysis should include multiple dilutions of highly reactive samples to better understand antibody concentrations (24).

It should be noted that baseline reactivity may or may not indicate prior exposure to EBOV – but could also be the result of, for example, previous exposure to a close relative of EBOV, possibly unknown and possibly non-pathogenic to humans. This association of pre-existing reactivity and odds of seroreactivity post-vaccination was not replicated in Beni, North Kivu Province. This may be due to geographically distinct exposure profiles of each cohort, where only 3.8% of Beni participants were considered EBOV-GP reactive at baseline, compared to 28.8% of participants in Mbandaka at baseline. The high baseline seroreactivity to EBOV GP in Mbandaka is not insignificant. This may be indicative of exposure to EBOV or a possible related pathogen circulating in the region, yet ultimately, we do observe increases in EBOV GP reactivity following vaccination. Further investigation into the observed background reactivity—whether EBOV GP specific or caused by exposure to another related antigen—is warranted.

Our results indicate similar trends in sustained anti-EBOV GP responses, yet differences in seroreactivity at the study site level. The odds ratios of seroreactivity in Beni were notably large, particularly at five-years post vaccination (OR = 9.28, 95% CI 4.45-19.35). This likely reflects the near complete seroconversion of all individuals included at this timepoint, which is supported by the 81% reactivity at Visit 7 with detected seroreactivity among this same cohort. These extreme odds may also be a product of the low baseline reactivity observed in Beni, as discussed. However, given the breadth of the associated confidence interval, there may be additional predictors not included in the model presented which could improve model fit and estimate precision. Future analysis should expand upon these mixed effects models to assess inclusion of alternative interaction terms or other indicators of exposure such as reactivity to other filovirus or non-filovirus antigens, underlying health conditions or possible co-infections with other pathogens, specific to the Beni site.

A decreased magnitude of anti-EBOV GP antibodies, and decreased odds of seroreactivity among male participants were observed across both sites, when compared to female vaccine recipients. These findings are aligned with published literature documenting differences in immune responses by sex broadly (25), where both anatomical and physiological differences as well as behavioral differences associated with gender roles may be driving these differences in observed antibody titers. For example, women may experience an increase in antibody magnitude and sustained seroreactivity over the five-year period due to their position as primary caretakers in the family ecosystem, and by extension increased exposure when providing care (26). These EBOV-specific findings are directly in line with previously published work from Beni at the six-month time point, where female sex was a significant predictor of seroreactivity (13).

Broadly, age was not identified as a significant predictor of seroreactivity or elevated antibody titers in this analysis. However, across both linear and logistic models, as participant age increased, we observed a trending decrease in the odds of seroreactivity, and antibody titer change relative to the eldest age group. This observation is supported by the well described notion of immunosenescence where the immune system undergoes age-related changes, which may affect vaccine-induced antibody durability (27). Furthermore, among elderly individuals, the immune system can be less responsive to vaccination (28), which supports our observed trend of reduced antibody titers with age among those recipients of the rVSV-ZEBOV-GP vaccine.

This study also spanned the administration of a second dose of the vaccine in Mbandaka at approximately 4.2 years (Visit 6) following initial deployment due to the recurrence of another recorded outbreak (29). The booster dose has been documented as well tolerated and inducing a robust response among a clinical trial participants in the US and Sierra Leone (9, 30), and our data similarly reflects a significant increase in the odds of seroreactivity and change in anti-EBOG GP antibody titers at Visit 6 and Visit 7 among the Mbandaka cohort. These data support the added value of booster dose administration, even to those who have a history of exposure whether by natural challenge with the virus or past vaccination. Alternative extended booster timelines have been suggested as an improved approach to promote antibody affinity maturation (31).

These facets of biological limits of antibody production and the timing of sample collection may also explain why we do not observe dramatic peaks of antibody reactivity to EBOV GP following documented recurrent EVD outbreaks at either site over the course of the five-year follow-up. Yet, it should not be underestimated that these recurrent documented outbreaks may be serving as natural booster opportunities to maintain anti-EBOV GP reactivity in individuals, even without documented EVD. Three individuals in this study population (one in Mbandaka and two in Beni) contracted and survived EVD within the study period during subsequent EBOV outbreaks. Serologically, this was further supported by evidence of elevated EBOV nucleoprotein (NP) and viral matrix protein 40 (VP40) (publication forthcoming). While this is a small sample, it serves as empirical anecdotal evidence of vaccine effectiveness with real-world challenge by the pathogen itself.

While anti-EBOV GP IgG is not a definitive correlate of protection, it remains one of the most widely used immunological markers in studies of EVD exposure and vaccine dynamics (12, 32). Our findings of sustained seroreactivity are consistent with prior reports from controlled cohorts in Europe and Central Africa, which have demonstrated persistence of GP-specific antibodies for up to five years post-vaccination (12, 32). In addition to understanding durability over a 5-year period, our cohort also reflects individuals vaccinated during active outbreaks, including frontline workers and contacts of confirmed cases, thereby providing critical insight into vaccine performance under real-world conditions in a filovirus endemic region such as the DRC.

This study has several limitations. First, the multiplex immunoassay approach to evaluating serologic responses does not provide an absolute titer, but instead magnitude of relative change in detected antibody responses. While this specific assay has been well characterized and compared with the gold-standard single plex FANG assay (16), all changes in antibody titers are relative to an established baseline at Visit 0 with reference to positive and negative controls. Second, loss to follow-up over the course of this five-year longitudinal study should not be discounted as a potential introduction of bias, yet overall retention over 60% in the DRC context is considered high. Furthermore, residual confounding by unmeasured exposure is likely, particularly in the DRC setting where ongoing transmission of EBOV and detection of seroreactivity to other antigens of the filovirus family has been documented (14, 33). The DRC had reported three sequential EVD outbreaks in Beni and two in Mbandaka over the course of this study follow up period, which could all contribute to the patchwork of exposures over time experienced by this study population. Given the inclusion of pan-filoviral targets, further analysis highlighting changes in this family-wide reactivity should be conducted. Lastly, these results solely report detected seroreactivity to EBOV GP, but do not specifically address the functionality of the reactive antibodies. While seroreactivity is a well-established measure of the ability of the immune system to detect the antigen presented, further investigation is needed to determine functional characteristics such as, for example, the neutralization capacity and avidity of these antibodies.

Additionally, further analysis can be conducted to not only determine magnitude change or odds of seroreactivity, but rather the durability of antibodies elicited by the rVSV-ZEBOV-GP vaccine. Future work should rely on survival analysis applied to the antibodies themselves to determine proportional hazards of antibody waning post-vaccination. In addition, EBOV NP and EBOV VP40 were included to confirm cases of natural infection as these antigens are not present in the rVSV-ZEBOV-GP vaccine. Similar analyses should be conducted to evaluate whether the detected changes in seroreactivity are specific to EBOV GP or also matched by increases in seroreactivity to EBOV NP and EBOV VP40, potentially indicating exposure to the wildtype virus, as opposed to the GP solely presented by the vaccine. Other filovirus targets should also be evaluated over time to ascertain potential exposure to other viruses of the same family in these regions, which may indicate sub-clinical infection or under-detection of outbreaks in the DRC setting.

Taken together, our results demonstrate the remarkable durability of anti-EBOV GP antibodies following vaccination with rVSV-ZEBOV-GP in an endemic setting. These results reinforce the value and efficacy of vaccination as a preventative approach to curb EVD outbreaks (10) and serve as real world evidence of sustained immune reactivity to the virus, particularly in ring vaccination frameworks (8). At the same time, the observed variability by age and geography suggests that targeted booster strategies—potentially prioritizing older individuals or high-risk occupational groups such as healthcare workers—may be warranted. Given the increasing frequency of EVD outbreaks in the DRC, and more broadly, the current Bundibugyo virus outbreak in May 2026 (33), proactive vaccination strategies for frontline workers, rather than reactive deployment alone should be considered for larger public health action (34).

## Data Availability

Data will be made available upon reasonable request to the corresponding author.

---

**Supplementary Figure 1.**
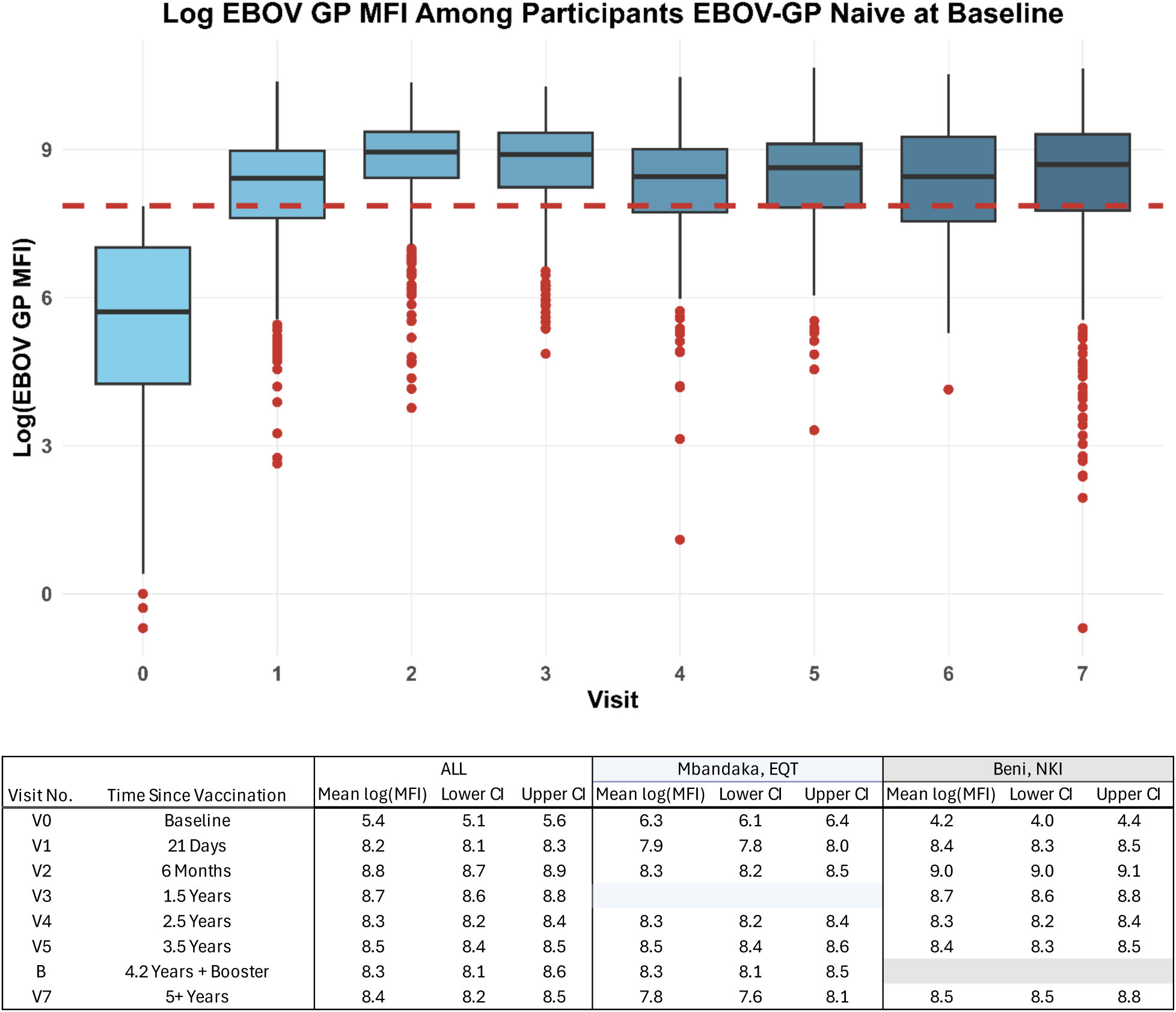
Mean Detected Log(MFI) across Visits, by Site among only those participants EBOV GP-naïve at baseline (n = 919).

**Supplementary Table 2.** Seroreactivity across visits among participants EBOV GP-naïve at baseline (n = 919).

| Covariate Strata | BASELINE<br>N | V1 |  | V2 |  | V3 |  | V4 |  | V5 |  | V6 |  | V7 |  |
| --- | --- | --- | --- | --- | --- | --- | --- | --- | --- | --- | --- | --- | --- | --- | --- |
|  |  | n | % | n | % | n | % | n | % | n | % | n | % | n | % |
| *Censor those Reactive at Baseline* |  |  |  |  |  |  |  |  |  |  |  |  |  |  |  |
| **Site** |  |  |  |  |  |  |  |  |  |  |  |  |  |  |  |
| Beni, NKI | 576 | 366 | 69.6 | 453 | 93.6 | 369 | 81.8 | 244 | 67.8 | 275 | 72.6 | -- | -- | 326 | 79.9 |
| Mbandaka, EQT | 343 | 141 | 42.5 | 182 | 61.5 | -- | -- | 150 | 60.7 | 151 | 66.5 | 80 | 56.7 | 111 | 51.5 |
| **Age Category** |  |  |  |  |  |  |  |  |  |  |  |  |  |  |  |
| 12-19 | 70 | 35 | 54.7 | 53 | 85.5 | 33 | 78.6 | 32 | 78.1 | 32 | 74.4 | 1 | 14.3 | 29 | 70.7 |
| 20-29 | 204 | 109 | 58.0 | 145 | 84.8 | 104 | 85.3 | 88 | 68.2 | 102 | 75.0 | 11 | 64.7 | 102 | 75.6 |
| 30-39 | 294 | 171 | 62.9 | 186 | 77.2 | 115 | 81.0 | 122 | 66.0 | 132 | 71.4 | 18 | 45.0 | 130 | 66.0 |
| 40-49 | 200 | 111 | 58.1 | 142 | 83.5 | 71 | 83.5 | 90 | 64.8 | 93 | 71.0 | 29 | 69.1 | 115 | 77.7 |
| 50+ | 151 | 81 | 56.6 | 109 | 80.2 | 46 | 76.7 | 62 | 54.9 | 67 | 60.4 | 21 | 60.0 | 61 | 58.7 |
| **Respondent Sex** |  |  |  |  |  |  |  |  |  |  |  |  |  |  |  |
| Male | 606 | 332 | 59.3 | 408 | 78.9 | 226 | 79.6 | 256 | 64.0 | 276 | 69.5 | 55 | 56.7 | 283 | 68.9 |
| Female | 313 | 175 | 58.7 | 227 | 86.3 | 143 | 85.6 | 138 | 66.7 | 150 | 71.8 | 25 | 56.8 | 154 | 72.0 |
| **Civil Status** |  |  |  |  |  |  |  |  |  |  |  |  |  |  |  |
| Single | 321 | 179 | 60.1 | 233 | 84.4 | 156 | 83.9 | 148 | 71.2 | 160 | 75.1 | 20 | 62.5 | 155 | 73.5 |
| Married or Living Together as Married | 571 | 315 | 58.9 | 382 | 79.4 | 203 | 80.2 | 235 | 61.5 | 257 | 67.8 | 58 | 55.8 | 271 | 68.1 |
| Divorced, Separated or Widowed | 24 | 12 | 54.6 | 17 | 85.0 | 8 | 80.0 | 9 | 60.0 | 9 | 64.3 | 2 | 40.0 | 9 | 64.3 |
| Unknown | 3 | 1 | 33.3 | 3 | 100.0 | 2 | 100.0 | 2 | 100.0 | -- | -- | -- | -- | 2 | 100.0 |
| **Educational Level** |  |  |  |  |  |  |  |  |  |  |  |  |  |  |  |
| No Formal Education | 28 | 11 | 55.0 | 17 | 100.0 | 12 | 80.0 | 7 | 50.0 | 6 | 42.9 | 0 | 0.0 | 8 | 61.5 |
| Any Primary School or Apprenticeship | 319 | 149 | 60.8 | 198 | 88.4 | 141 | 80.6 | 102 | 63.4 | 123 | 72.8 | 8 | 42.1 | 131 | 72.0 |
| Completed Secondary School | 291 | 138 | 61.6 | 166 | 78.7 | 94 | 81.7 | 97 | 62.2 | 106 | 65.8 | 23 | 60.5 | 119 | 73.9 |
| Post-Secondary Education | 442 | 248 | 67.4 | 277 | 84.7 | 124 | 84.9 | 204 | 74.2 | 207 | 79.0 | 59 | 71.1 | 189 | 70.5 |
| Refused | 1 | 0 | 0.0 | 1 | 100.0 | -- | -- | 1 | 100.0 | -- | -- | -- | -- | 1 | 100.0 |
| **Occupation (by Exposure Route)** |  |  |  |  |  |  |  |  |  |  |  |  |  |  |  |
| Health Care Worker | 319 | 207 | 67.2 | 224 | 80.0 | 132 | 82.5 | 151 | 66.2 | 150 | 67.2 | 44 | 68.8 | 169 | 71.9 |
| Zoonotic Exposure (Farmers, Hunters) | 173 | 87 | 55.8 | 117 | 83.0 | 75 | 80.7 | 64 | 58.2 | 70 | 60.9 | 7 | 41.2 | 76 | 66.1 |
| Human to Human Exposure (Merchants, Teachers, Politicians) | 126 | 54 | 46.6 | 83 | 78.3 | 52 | 86.7 | 46 | 61.3 | 54 | 71.1 | 9 | 42.9 | 54 | 68.4 |
| Other | 293 | 157 | 57.9 | 204 | 82.9 | 106 | 79.7 | 127 | 67.9 | 149 | 79.2 | 20 | 51.3 | 132 | 69.5 |
| Unknown, Refused | 8 | 2 | 28.6 | 7 | 100.0 | 4 | 80.0 | 6 | 85.7 | 3 | 75.0 | -- | -- | 6 | 100.0 |

